# Effectiveness and cost-effectiveness of public health measures to control COVID-19: a modelling study

**DOI:** 10.1101/2020.03.20.20039644

**Authors:** Qiang Wang, Naiyang Shi, Jinxin Huang, Tingting Cui, Liuqing Yang, Jing Ai, Hong Ji, Ke Xu, Tauseef Ahmad, Changjun Bao, Hui Jin

## Abstract

**Background:** The Severe Acute Respiratory Syndrome Coronavirus 2 (SARS-CoV-2) was first reported in China, which caused a respiratory disease known as Coronavirus Disease 2019 (COVID-19). Since its discovery, the virus has spread to over 160 countries and claimed more than 9800 deaths. This study aimed to assess the effectiveness and cost-effectiveness of various response public health measures.

**Methods:** The stochastic agent-based model was used to simulate the process of COVID-19 outbreak in scenario I (imported one case) and II (imported four cases) with a series of public health measures, involving the personal protection, isolation-and-quarantine, gathering restriction, and community containment. The virtual community was constructed following the susceptible-latent-infectious-recovered framework. The epidemiological and economic parameters derived from the previous literature and field investigation. The main outcomes included avoided infectors, cost-effectiveness ratios (CERs), and incremental cost-effectiveness ratios (ICERs). The sensitivity analyses were undertaken to assess uncertainty.

**Results:** In scenario I and II, the isolation-and-quarantine averted 1696 and 1990 humans infected respectively at the cost of US$12 428 and US$58 555, both with negative value of ICERs. The joint strategy of personal protection and isolation-and-quarantine could avert one more case than single isolation-and-quarantine with additional cost of US$166 871 and US$180 140 respectively. The effectiveness of isolation-and-quarantine decreased as lowering quarantine probability and increasing delay-time. Especially in scenario II, when the quarantine probability was less than 25%, the number of infections raised sharply; when the quarantine delay-time reached six days, more than a quarter of individuals would be infected in the community. The strategy including community containment could protect more lives and was cost-effective, when the number of imported cases was no less than 65, or the delay-time of quarantine was more than five days, or the quarantine probability was below 25%, based on current assumptions.

**Conclusions:** The isolation-and-quarantine was the most cost-effective intervention. However, personal protection and isolation-and-quarantine was the optimal strategy averting more infectors than single isolation-and-quarantine. Certain restrictions should be considered, such as more initial imported cases, longer quarantine delay-time and lower quarantine probability.

## Introduction

As of March 21, 2020, about 81 008 cases of coronavirus disease 2019 (COVID-19) have been identified in China [1]. The global number of reported cases of COVID-19 has surpassed 230 000 and the confirmed cases of COVID-19 have been reported in more than 160 countries [2]. As date, the 21th century has witnessed several large-scale outbreaks of infectious diseases caused by coronaviruses. The cases infected with COVID-19 were significantly higher than ones infected with Severe Acute Respiratory Syndrome (SARS) and Middle East Respiratory Syndrome (MERS). The statistic showed that SARS caused more than 8000 morbidity, while MERS with more than 2200 morbidity involving over 25 countries worldwide [3].

The outbreak of COVID-19 posed an emergency of international concern and challenges in the absence of specific therapeutic treatment and vaccine. On March 11, 2020 COVID-19 has been declared a pandemic by the World Health Organization (WHO). The nonpharmaceutical interventions (NPIs) are necessary to prevent person-to-person transmission of COVID-19. So far, a series of measurements have been implemented in China. The isolation of infected cases and quarantine of humans exposed to the cases were the most common public health measures. The susceptible humans wear masks and maintain good hygiene practices. The Chinese authorities have introduced restrictions on public gathering, unnecessary movements, and public transportation.

The success of NPIs depended on the epidemiological characteristics of the disease as well as the effectiveness of the measures [4]. The previous studies suggested that the effectiveness of interventions was associated with the proportion of asymptomatic infection, transmissibility of the virus, and the intervention feasibility [4, 5].

Hence, we need specific, effective, and cost-effective guidance framework of measures to deal with emerging COVID-19 in community transmission. The strategy framework was need to be developed based on the epidemiological characteristics, intervention feasibility, and economic cost.

The purpose of this study was to assess the effectiveness and cost-effectiveness of different public health measures and provide the suggestions and assist decision and policy makers in making better decisions and resources allocations in the fight with COVID-19 outbreak.

## Method

### Model

The stochastic agent-based model (ABM) was used to simulate the process of COVID-19 outbreak with different interventions. The Netlogo software was applied to run the simulation. We constructed the domain with a total number of agents, following the susceptible-latent-infectious-recovered (SLIR) framework. In the domain, each agent was susceptible initially, then the COVID-19 cases were introduced into the agents. The infectious agent could infect the susceptible agents with the infectious ability following the distance transmission probability function of *β* (*r*) [6]. The simulation would stop when there did not exist an exposed or infected agent in the space. We assumed that the recovered agents would not become susceptible again. One or four COVID-19 cases were introduced randomly locating on the simulated space with 2000 humans separately as two scenarios, representing sporadic (one imported case) and cluster (four imported case) of outbreak respectively.

### Comparator strategies

In the study, we involved the measures including personal protection, isolation-and-quarantine, gathering restriction, and community containment. We combined these measures to form various joint intervention strategies. Four joint interventions were formulated: program A: personal protection and isolation-and-quarantine, program B: gathering restriction and isolation-and-quarantine, program C: personal protection and community containment, program D: personal protection, isolation-and-quarantine, and gathering restriction. We compared different single and joint strategies versus no-interventions.

The systematic reviews have proved that it was useful to reduce the transmission of respiratory viruses with personal physical interventions [7–9]. In our study, we defined personal protection as both mask wearing and frequent hand washing. The isolation referred to the isolation of the symptomatic infected individuals, and quarantine referred to the tracing and quarantine of close contacts of symptomatic infected for a certain time [4, 10]. These practical tools have been going on for hundreds of years in the fight of infectious disease [11]. In previous outbreaks such as SARS in 2003 and Ebola in 2014 controlling the spread of infectious diseases has been proven by using isolation and quarantine [12–15].

The gather restriction and community containment belonged to the social distancing which was designed to reduce personal interactions and thereby transmission risks [16]. In China, restrictions on gathering referred to the restriction of crowd-gathering activities, especially catering and entertainment. The enforcement of community containment was a restriction on the movement of people throughout the community, minimizing human contact [17].

### Epidemiological parameter

The incubation period and serial interval came from the estimation of Chinese Center for Disease Control and Prevention (CDC) and Guangdong Provincial CDC in the field work [18, 19], and were considered fitting to the gamma distribution in the model [3]. The parameter of distance transmission probability has been reported in previous study [6]. The protective effectiveness of personal physical interventions derived from the cluster randomized controlled trial [20]. In our study, we converted odds ratio (OR) of handwashing and mask-wearing into the relative risk (RR), and calculated the (1-RR)/RR as the personal protection effectiveness [21].

In the model, we set the probability and delay-time for isolation and quarantine. The isolation delay-time meant that the time of dealing with patients lagged behind the time of infection onset, and the quarantine delay-time meant that the time of handling close contact lagged behind the time of exposing. Initially, we assumed that the index case (initial imported case) would be 100% isolated with no time delay (infecting others and isolation were carried out within the same day and infecting others preceded isolation). The quarantine probability was 100% and delay-time was two days. In the sensitivity analysis, the probability of quarantine of close contacts was set from 25% to 100% and the delay-time was from zero day to six days.

### Cost

The economic data derived from the field work and previous literature (Table 1). The cost of personal protection included masks and handwashing (water and soap). The price of the mask was US$0.14 each and we assumed that two masks were used per person per day [22]. Given the soap using, the cost of handwashing per person per day was calculated as the formula provided in the previous study [23]:

**Table 1.**
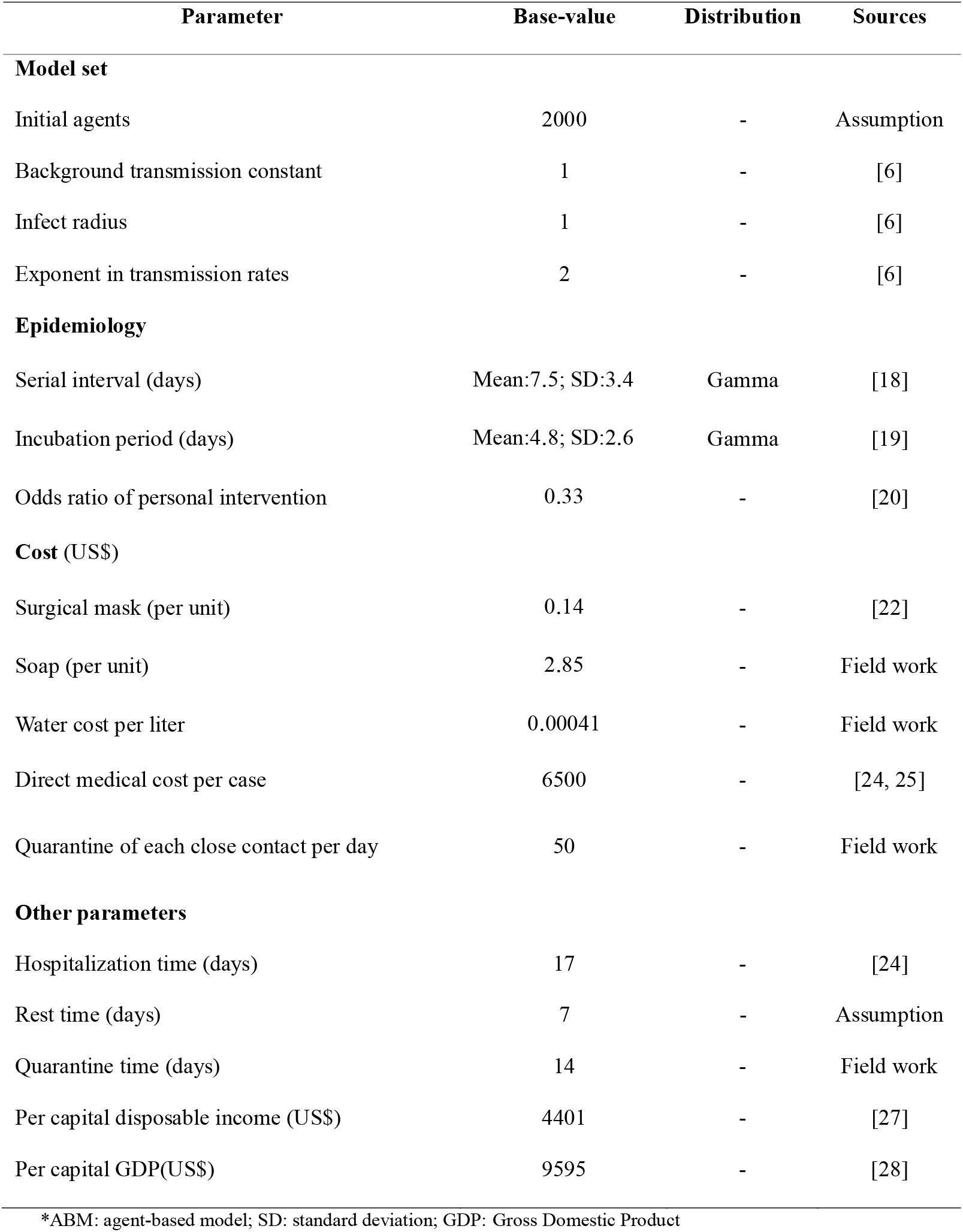
Parameters in the ABM model

| Parameter | Base-value | Distribution | Sources |
| --- | --- | --- | --- |
| **Model set** |  |  |  |
| Initial agents | 2000 | - | Assumption |
| Background transmission constant | 1 | - | [6] |
| Infect radius | 1 | - | [6] |
| Exponent in transmission rates | 2 | - | [6] |
| **Epidemiology** |  |  |  |
| Serial interval (days) | Mean:7.5; SD:3.4 | Gamma | [18] |
| Incubation period (days) | Mean:4.8; SD:2.6 | Gamma | [19] |
| Odds ratio of personal intervention | 0.33 | - | [20] |
| **Cost (US\$)** | | | |
| Surgical mask (per unit) | 0.14 | - | [22] |
| Soap (per unit) | 2.85 | - | Field work |
| Water cost per liter | 0.00041 | - | Field work |
| Direct medical cost per case | 6500 | - | [24, 25] |
| Quarantine of each close contact per day | 50 | - | Field work |
| **Other parameters** |  |  |  |
| Hospitalization time (days) | 17 | - | [24] |
| Rest time (days) | 7 | - | Assumption |
| Quarantine time (days) | 14 | - | Field work |
| Per capital disposable income (US\$) | 4401 | - | [27] |
| Per capital GDP(US\$) | 9595 | - | [28] |

\*ABM: agent-based model; SD: standard deviation; GDP: Gross Domestic Product

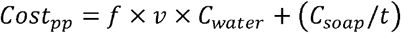

where the cost_pp_ = cost of hand washing, f= times of hand washing per day, and we set to six, v= volume of hand washing per time, and we set to 1000c.c/ml, C_water_ = water cost per liter, and was US$0.00041, C_soap_ = cost of soap, and was US$2.85, t = the number of days soap available, and we set to 60. We assumed the day of personal protection was equal to the time from the day first case occurred to the last case recovered in the area plus 14 days.

The cost of cases included the direct medical cost and indirect cost. We searched the cost of SARS patients to estimate the COVID-19 cases. In Guangzhou, China, the average hospitalization cost per patient was US$2900, and the average hospital stay was 17 days [24]. The average hospitalization cost achieved US$10 000 in Canada [25], which was higher than that in China [24]. We estimated the average medical cost of US$6500 for COVID-19 patient. Referring to human capital approach in disease burden [26], we estimate that the indirect cost of infected patient using per capita disposable income (PCDI)/365.25* (hospitalization days added rest days). The average rest days were estimated to seven days. We assumed that the cost of isolation would be included in the cost of hospitalization. The cost of quarantine of close contacts included direct and indirect parts. The cost of quarantine (accommodation and surveillance daily) per day was US$50 for each close contact. Similar to human capital approach in disease burden [26], the indirect cost of quarantine of close contact was calculated through PCDI/365.25* days of quarantine. For COVID-19, the quarantine time of close contact was about 14 days in China. There was very limited evidence providing the estimation method of cost of gathering restriction and community containment. Referring to the human capital, we posed the following formulas to roughly estimate the cost of gathering restriction community containment during current COVID-19 outbreak in China.

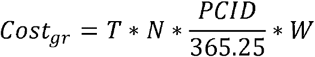

where the cost_gr_ = cost of gathering restriction, T= days of restriction, N= total number of humans in the space, PCID = per capita disposable income, W: weight of calculation, and we set to 0.3.

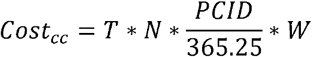

where the cost_cc_ = cost of community containment, T= days of containment, N= total number of humans in the space, PCID = per capita disposable income, W: weight of calculation, and we set to 0.8. We assumed the day of restriction and containment equal to the time from the day first case occurred to the last case recovered in the region plus 14 days. We used PCDI in 2018 China deriving from the National Bureau of Statistical [27]. In our study, all costs (RMB) were converted into US$ as per 2018 currency conversion rate available from (http://www.safe.gov.cn/safe/2018/0928/10272.html) (1 US$=6.879 RMB).

### Measurement of cost-effectiveness

Main health benefits of our study were avoided infections conducting measures versus no-interventions. The cost-effectiveness ratios (CERs) and incremental cost-effectiveness ratios (ICERs) were calculated as the main cost-effectiveness outcomes. We calculated the CERs for interventions through cost divided by humans protected (uninfected). The ICERs were calculated as the difference in the total costs between the intervention cohorts and non-intervention cohorts, divided by the difference in the total avoided infection. Positive ICERs showed the incremental costs required for avoiding 1 infected person. Negative ICERs indicated that intervention results in fewer costs while avoiding infected people than no intervention. The strategy was considered to be cost-effective if ICERs were lower than three times of per capita GDP. In 2018, the per capita GDP in China was US$9595 [28]. We did not discount the cost because of the short time span of the analysis. We performed the 1000 Monte Carlo simulations and reported the mean and standard deviation of the result of runs. Reporting of methods and results conformed to the Consolidated Health Economic Evaluation Reporting Standards (Additional file 1: Table S1) [29].

One-and-two-way sensitivity analyses were performed to explore impact of the parameters in the range to test the robustness of the findings, including the epidemiological characteristics, interventions implement, and economic parameters.

## Results

### Effectiveness of measures in scenario I (one case)

Introduction of one case, each strategy could avoid the number of infectors and be cost-effective compared with no intervention (Table 2). The isolation-and-quarantine was the most cost-effective intervention, avoiding 1696 cases and saving US$11 515 944 (ICERs < 0). The most protective single strategy was community containment, which avoided one more case than the isolation-and-quarantine at the additional US$549 186. Among the joint strategies, there was the lowest ratio of cost-effectiveness for the program A (CERs= 90 US$/ per human protected). The program A could avert one more infector comparing to single isolation-and-quarantine.

**Table 2.** The cost-effectiveness of intervention measures in scenario I (US$1 000)

| Type | Intervention strategy | Infectors | Cost |  |  | CERs | ICERs |
| --- | --- | --- | --- | --- | --- | --- | --- |
|  |  |  | Cost of cases | Cost of measures | Total cost |  |  |
|  | No intervention | 1698±716.41 | 11 528.37±4863.86 | 0 | 11 528.37±4863.86 | - | - |
| Single | Personal protection | 1319±950.02 | 8952.90±6449.89 | 486.97±260.28 | 9439.86±6700.97 | 13.856 | -5.505 |
|  | Isolation-and-Quarantine | 2±1.08 | 10.46±7.38 | 1.97±1.55 | 12.43±8.40 | 0.006 | -6.788 |
|  | Gathering restriction | 1595±800.23 | 10 825.76±5432.90 | 1155.65±512.30 | 11 981.41±5924.80 | 29.552 | 4.378 |
|  | Community containment | 1±0.70 | 9.64±4.75 | 551.97±52.19 | 561.61±53.21 | 0.281 | -6.464 |
| Joint | Program A | 1±0.47 | 9.23±4.58 | 170.07±22.89 | 179.30±25.04 | 0.090 | -6.690 |
|  | Program B | 1±0.72 | 9.44±3.46 | 226.73±34.45 | 236.17±37.23 | 0.118 | -6.656 |
|  | Program C | 1±0.48 | 8.55±3.29 | 712.60±60.02 | 721.15±60.79 | 0.361 | -6.369 |
|  | Program D | 1±0.75 | 8.83±5.06 | 403.11±63.42 | 411.94±66.87 | 0.206 | -6.552 |

\* Program A: personal protection and isolation-and-quarantine; Program B: isolation-and-quarantine and gathering restriction; Program C: personal protection and community containment; Program D: personal protection, isolation-and-quarantine, and gathering restriction; CERs: cost-effectiveness ratios; ICERs (incremental cost-effectiveness ratios): compared with no intervention.

### Effectiveness of measures in scenario II (four cases)

In scenario II (Table 3), compared with no intervention, personal protection or gathering restriction was not cost-effectiveness (ICERs > three times of per capita GDP). The isolation-and-quarantine was still the most cost-effective, avoiding 1990 cases and saving US$13 372 397 (ICERs< 0). Compared with the isolation-and-quarantine, community containment could avoid one more case with the additional US$600 044. Among the joint strategies, there was the lowest ratio of cost effectiveness for the program A (CERs= 121 US$/ per human saved). Similarly, the program A versus single isolation-and-quarantine could avert one more infector.

**Table 3.**
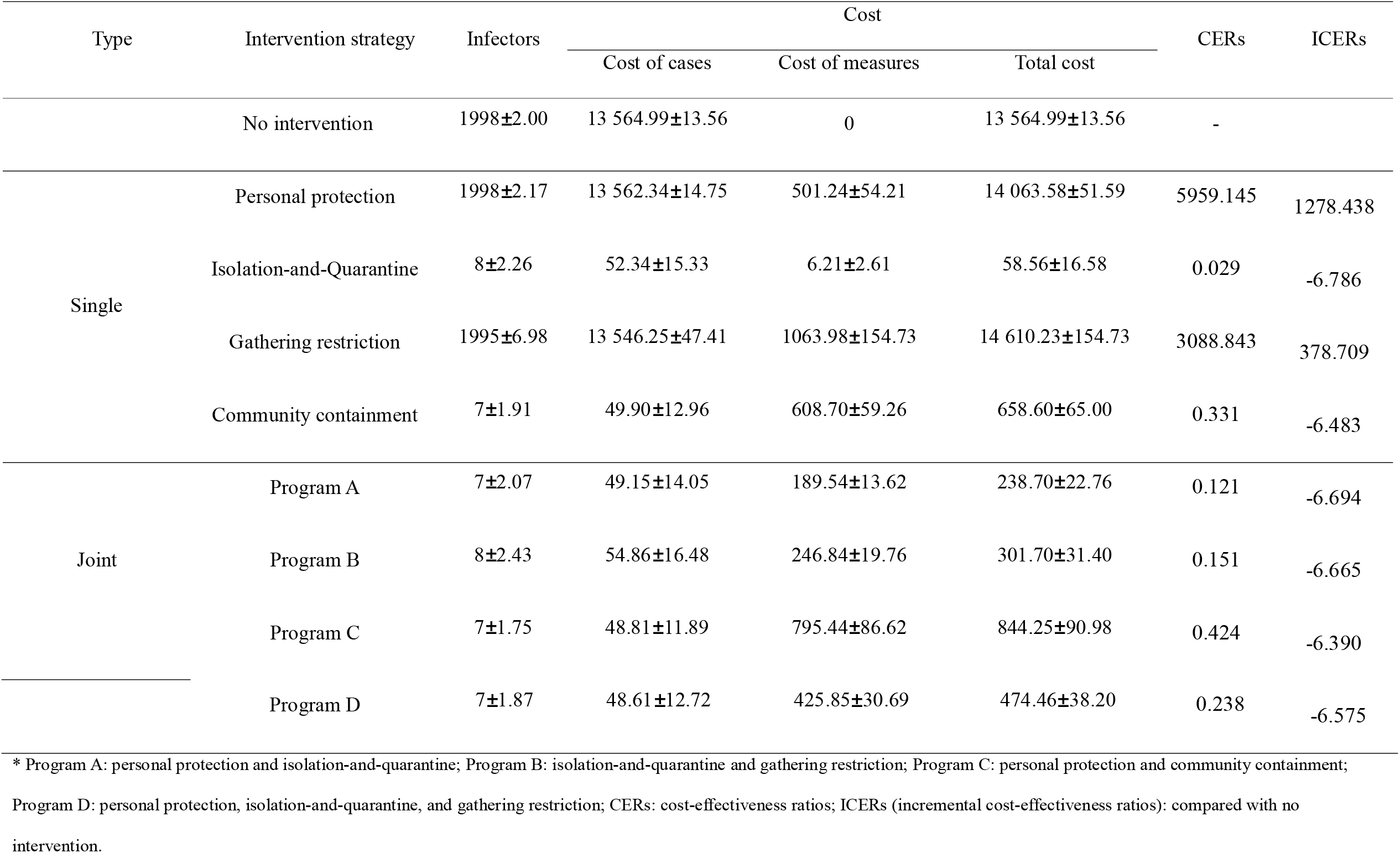
The cost-effectiveness of intervention measures in scenario II (US$1 000)

### One-way sensitivity analysis

#### Transmission constant

The number of infectors depended on transmission constant in scenario I (Additional file 1: Table S2), while it kept stable after changing transmission constant in scenario II (Additional file 1: Table S3). The infectors obviously increased as the rise of transmission constant in scenario I. The effective reproduction number (Re) was 1.84 (95%CI: 1.81, 1.87) and 3.80 (95%CI: 3.53, 4.06) respectively when the transmission constant was one in scenario I and II (Additional file 1: Table S4). Varying the transmission constant from the 0.25 to two, the isolation-and-quarantine was the most cost-effective single intervention, and program A was the most cost-effective joint intervention.

#### Initial introduced cases

The number of imported cases was a key parameter influencing the effectiveness and cost-effectiveness analysis. There were not significantly differences in effectiveness between the program A and C, when the imported cases were set to ten or 20 (Figure 1a and Additional file 1: Table S5). When the imported cases were no less than 50, the program C including community containment could effectively decrease the infectors than program A including isolation-and-quarantine, but the former was not cost-effective. The CERs of interventions increased significantly as the increase of imported cases (Figure 2a). The threshold analysis showed that program C became cost-effective (ICERs< three times of per capita GDP) comparing to program A when initial cases increased to imported 65 cases (Additional file 1: Table S6).

**Figure 1.**
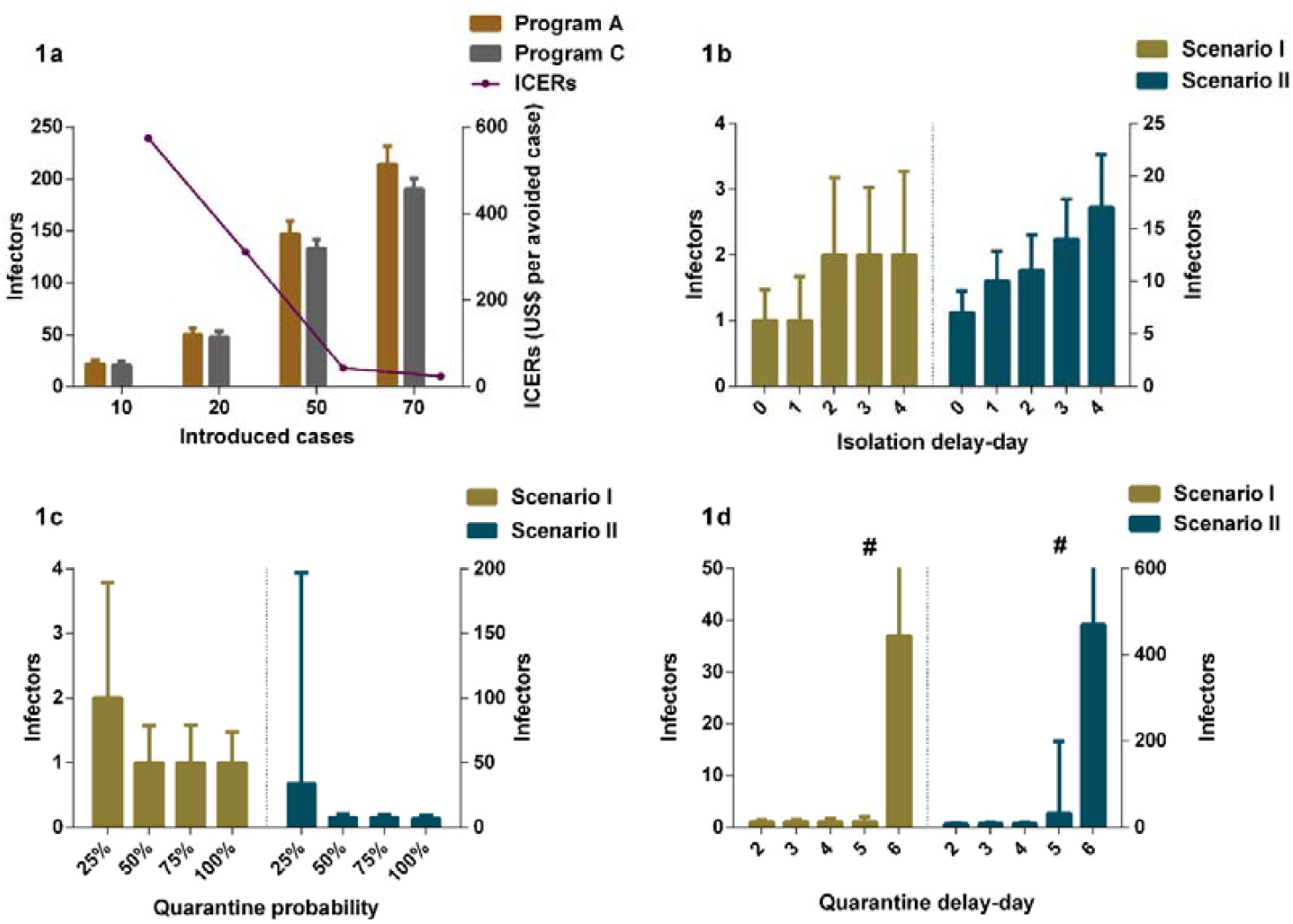
The impact of different parameters on interventions effectiveness. 1a: The comparisons of infections in program A and C in different introduced cases; 1b: The impact of isolation delay-day in program A in different scenarios; 1c: The impact of quarantine probability in program A in different scenarios; 1d: The impact of quarantine delay-day in program A in different scenarios; Program A: personal protection and isolation-and-quarantine; Program C: personal protection and community containment. ICERs: Incremental cost-effectiveness ratios # The interval extends out of the plotting region.

**Figure 2.**
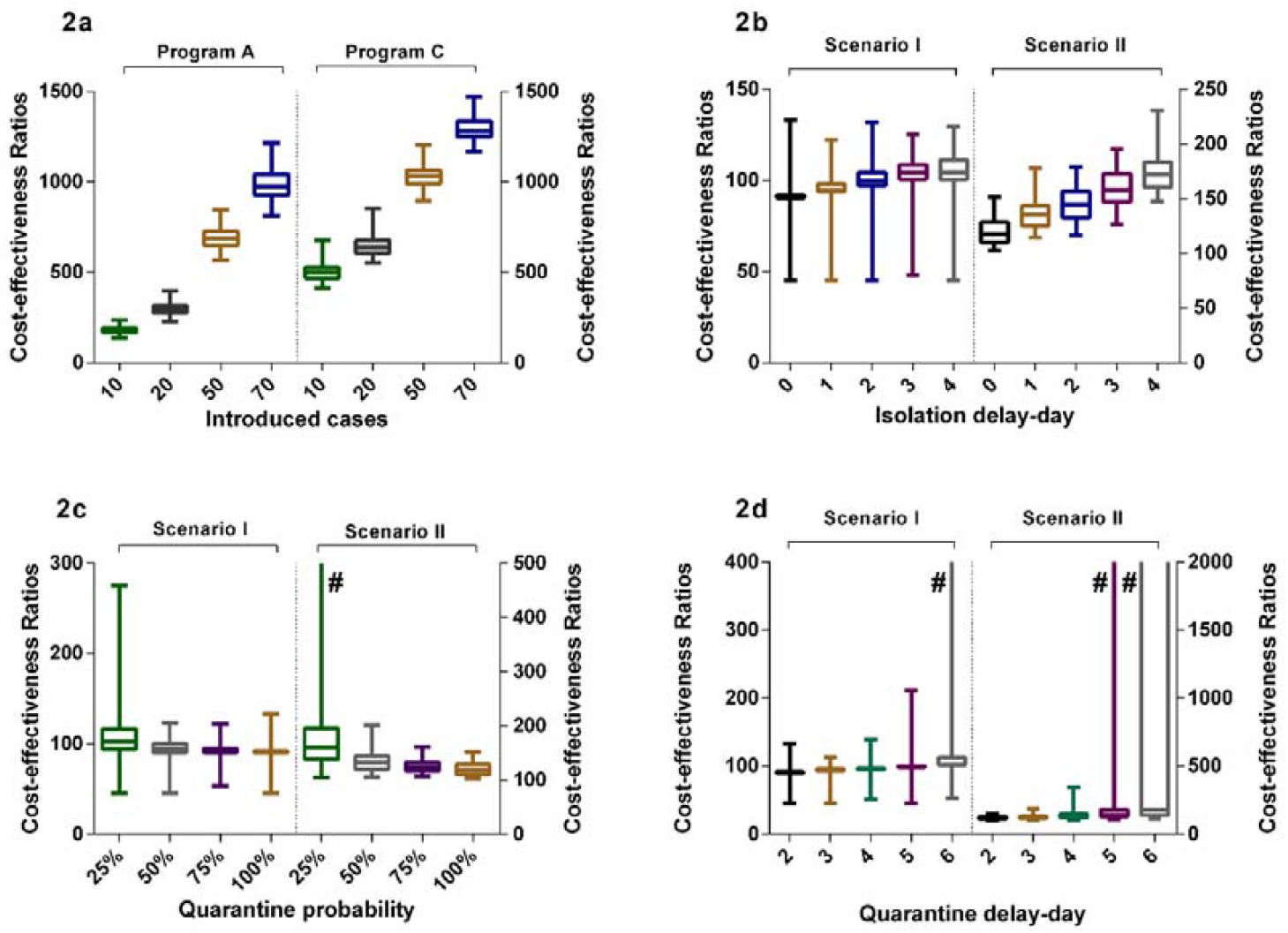
The Cost-effectiveness ratios of interventions with different parameters. 2a: The cost-effectiveness ratios in program A and C in different introduced cases; 2b: The cost-effectiveness ratios of different isolation delay-day in program A in different scenarios; 2c: The cost-effectiveness ratios of different quarantine probability in program A in different scenarios; 2d: The cost-effectiveness ratios of different quarantine delay-day in program A in different scenarios; Program A: personal protection and isolation-and-quarantine; Program C: personal protection and community containment. # The box or interval extends out of the plotting region.

#### Isolation delay-time

The isolation delay-time did not contribute to the spread of infections in scenario I (Figure 1b). The increase of isolation delay time, however, caused a significant increase in the number of infections in scenario II. When the isolation delay of four index cases reached four days, there were more than 15 humans being infected, which was three times as the one without isolation delay. The CERs of interventions increased as the increase of the isolation delay-day (Figure 2b). The program A dominated the program C in scenario I and II within the sensitivity analysis of isolation delay-time (Additional file 1: Table S7).

#### Quarantine probability

The effectiveness of isolation-and-quarantine was sensitive to the low quarantine probability. When the tracing probability of close contact was reduced to 25%, the number of people infected increased significantly, especially in the scenario II (Figure 1c). In scenario I and II, the effectiveness of outbreak controlling was close between program A and C when the probability of tracing above 50% (appendix table 8 and table 9). The CERs decreased as the increase of quarantine probability, and was most unstable when the quarantine probability was 25% (Figure 2c). In scenario I, the program C was not cost-effective comparing to program A. The ICERs of program C was close to three times of per capita GDP when the quarantine probability was 25% in scenario II. The threshold analysis showed that program C became cost-effective (ICERs< three times of per capita GDP) comparing to program A when quarantine probability was below 28% (Additional file 1: Table S10).

#### Quarantine delay-time

Varying the quarantine delay time from zero day to four days, it had little influence on averting infected cases (Figure 1d). When the tracing delay-time of close contacts was extended to six days, the number of people infected increased significantly (Additional file 1: Table S11 and Table S12). In scenario II, when quarantine delay-time reached six days, there were likely more than 500 humans being infected, accounting for a quarter in the space. The CERs of interventions was unstable when the quarantine delay-time was no less than five days (Figure 2d). Comparing with program A, the program C was cost-effective when the delay-time more than five days in scenario I and four days in scenario II respectively (ICERs< three times of per capita GDP).

#### Cost of patients

Varying the cost of patient from US$2900 to US$10 000, the CERs of interventions increased and ICERs of interventions comparing to the non-intervention decreased (Additional file 1: Table S13 and Table S14). The most cost-effective strategy was isolation-and-quarantine in scenario I and II.

### Two-way sensitivity analysis

#### Transmission constant and quarantine probability

In scenario I, the effectiveness of outbreak controlling was not sensitive to the transmission constant and quarantine probability (Additional file 1: Table S15). When the transmission constant was set to two, the outbreak could be controlled by the 25% probability quarantine. However, as the transmission constant increased in scenario II, the control of outbreak required higher quarantine probability. When the quarantine probability was 25% and transmission constant was two, it was likely about a quarter of people would be infected in scenario II (Additional file 1: Table S16). The program A dominated the program C in the scenario I and II in general. When the transmission constant was above one and the quarantine probability was below than 25%, the program C was cost-effective (ICERs< 3 times of per capita GDP).

#### Isolation delay-time and quarantine probability

In scenario I, the quarantine probability and isolation delay-time in the range of our analysis did not have a big effect on the cost-effectiveness results. However, when the quarantine probability was 25% and the isolation delay time reached three or four days, the variability of the effect of infection control increased (Figure 3a and Additional file 1: Table S17). In scenario II, the infectors increased significantly as the decrease of quarantine probability and increase of isolation delay-time (Figure 3b and Additional file 1: Table S18). In scenario II, when the quarantine probability decreased to 25%, the program C comparing to the program A was cost-effective (ICERs < three times of per capita GDP). When probability reached 50%, the program C would be cost-effective at the isolation delay-time more than two days.

**Figure 3.**
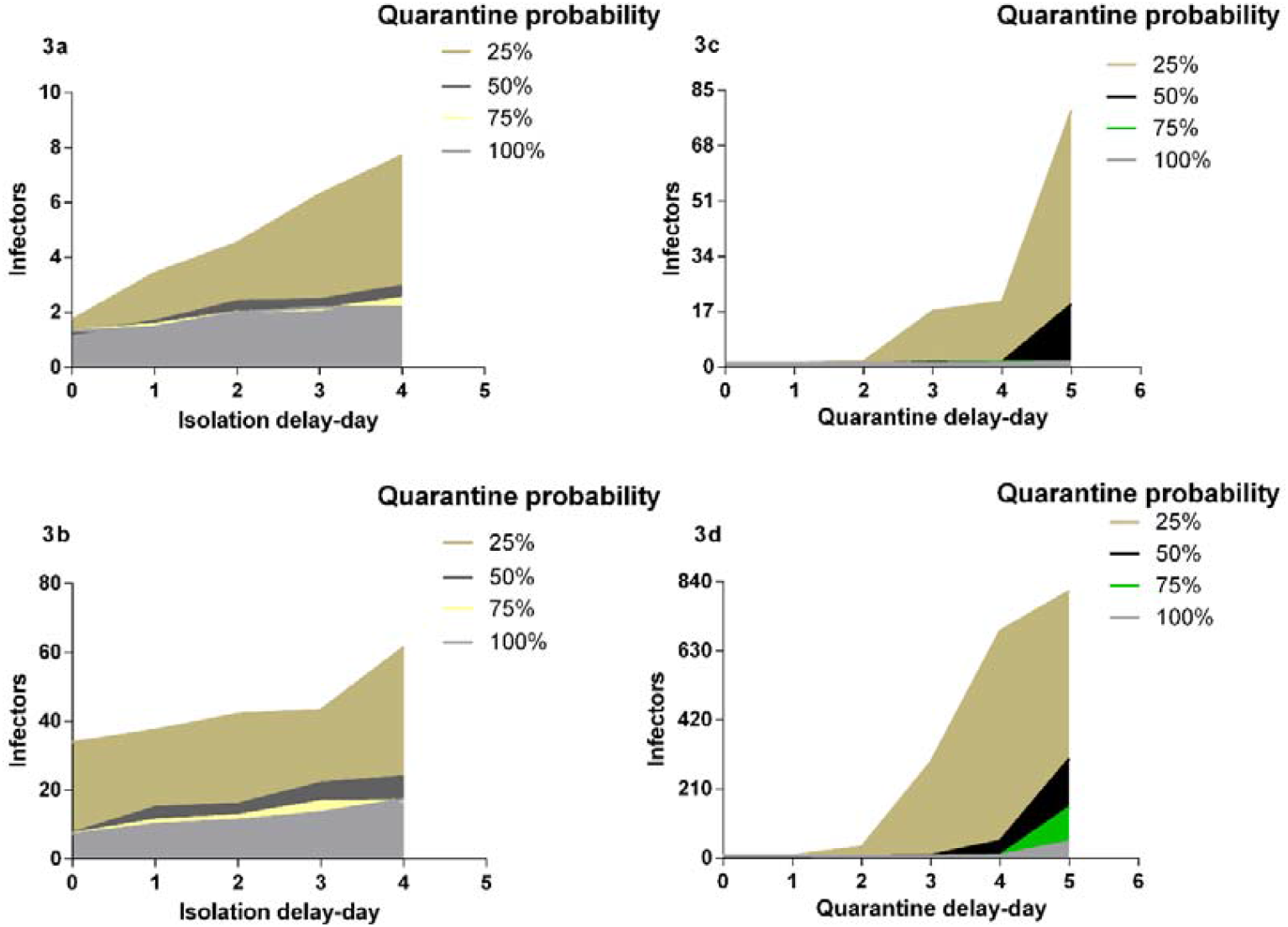
Impact of isolation and quarantine parameters on interventions effectiveness. 3a: the impact of isolation delay-time and quarantine probability in scenario I; 3b: the impact of isolation delay-time and quarantine probability in scenario II; 3c: the impact of quarantine delay-time and quarantine probability in scenario I; 3d: the impact of quarantine delay-time and quarantine probability in scenario II; Program A: personal protection and isolation-and-quarantine.

#### Quarantine delay-time and quarantine probability

The low quarantine probability and long quarantine delay-time would contribute to the outbreak of COVID-19, especially in scenario II (Figure 3c and 3d). At the 25% probability and five days’ delay-time, there were more than 800 humans infected in scenario II. The infectors increased significantly as the decrease of quarantine probability and increase of isolation delay-time. In scenario I, the program C was cost-effective comparing to the program A when the probability reached 25% and delay-time was more than two days, or when the probability reached 50% and delaying-time was more than four days (Additional file 1: Table S19). In scenario II, the program C was cost-effective when the probability reached 25%, or when the probability reached 50% and delaying-time was more than three days (Additional file 1: Table S20)

#### Cost parameter and quarantine probability

As increase of the cost of quarantine per close contact per day, the CERs of program A and program C increased but the ICERs of program C comparing to program A decreased. In scenario I, program A dominated the program C (Additional file 1: Table S21). In scenario II, the program C was cost-effective at the 25% quarantine probability. The CERs of program C were associated with the cost of community containment. Compared with program A, the CERs and ICERs of program C increased as the increase of cost of community containment (Additional file 1: Table S22). However, the optimal strategy was not affected by the cost of community containment.

#### Cost parameter and quarantine delay-time

Similarly, the cost of quarantine and cost of community containment were considered in the analyses. The both cost parameters had an effect on CERs, which increased as the increase of cost of quarantine and cost of community containment (Additional file 1: Table S23 and Table S24). The change of cost parameter did not have effect on the choice of optimal strategy.

## Discussion

Our study provided the assessment of different measures to control community transmission of COVID-19. In the sporadic (one imported case) and cluster (four imported cases) outbreak area, the isolation of infectious cases, and quarantine of humans exposing to the infections were the most cost-effective measure. In the virtual environment, isolation-and-quarantine could greatly reduce the number of infections and avoid the outbreak of disease with less cost. From the perspective of effectiveness and cost-effectiveness of controlling the spread of COVID-19, the joint strategy personal protection and isolation-and-quarantine was the optimal choice, averting more cases than single isolation-and-quarantine.

In the sporadic area, the effectiveness of isolation-and-quarantine was most sensitive to the quarantine delay-time. The one-way analysis showed that there was a marked increase in the number of infections when the quarantine delay-time reached six days. There were not significantly different about the number in the sporadic area when quarantine probability changed ranging from 25% to 100%. However, two-way analysis suggested that at the 25% probability, there were likely more infectors occurred when the quarantine delay-time was more than two days. In the cluster area, these parameters played an important role on the effectiveness of interventions. The probability of contact tracing decreased and delay-time of isolation-and-quarantine increased, leading to the fewer cases averted by the intervention. The long delay-time and low quarantine probability could accelerate the outbreak of COVID-19.

The effectiveness and cost-effectiveness of interventions were sensitive to the initial imported cases. The increase of imported cases could lead to the increase of risk of COVID-19 infection, even conducting the strict interventions. We suggested that the infectors avoided by isolation-and-quarantine and community containment were not significantly when the imported the cases below 20. When the imported cases reached 50, community containment could avoid more cases significantly. The strategy including community containment was cost-effective when imported cases reached 65, the 3.25% of the community population (2000 humans). The current article found that the initial number of cases had an effect on the effectiveness of interventions [30].

The choice of optimal strategy depended on the setting parameter of interventions. We compared the strategy of personal protection and isolation-and-quarantine (program A) with strategy of personal protection and community containment (program C). Generally, program A was cost-effective versus program C. however, the program C was cost-effective at the 25% probability and more than two quarantine delay-days, or 50% probability and no less than five quarantine delay-days in the sporadic outbreak area. The program C would dominate the program A at the 25% quarantine probability or quarantine delay-time was more than three days in the cluster area.

The effectiveness of isolation and contact tracing was associated with the extent of transmission before symptom onset [31]. The proportion of asymptomatic infection would contribute to the outbreak of COVID-19 [30], which was consistent with our findings. In our study, the community containment would be more efficient and cost-effective when the quarantine delay-time was more than latent period. We suggested that increase of quarantine-time delay was similar to the presence of asymptomatic infection. For asymptomatic infection or latent infection, failure to detect in time lead to the absence of isolation and continuation of transmission. We suggested that this phenomenon was similar to the low probability of quarantine or quarantine delay-time, which caused the infection to continue to spread. The proportion of asymptomatic infection had a big effect on the choice of controlling strategy.

There were some limitations in the study. First, COVID-19 was recently emerged disease first reported in Wuhan, China, therefore the availability of epidemiological data is insufficient. We set the study parameters referring to the existing published epidemiological studies and adopted the Gamma distribution to some of the parameters, which could improve the precision of estimate. Second, the cost of societal interventions was difficult to estimate. In our study, human capital approach was borrowed which might more conservatively estimate the cost. The cost of the disease would also increase, if according to the actual situation in Wuhan, China. Third, our model simulated a local area with 2000 humans, which may result in limited extrapolation ability. Finally, the simplification of the model will have some biases compared with the real situation, because the flow of people will be affected by many factors.

## Conclusion

In the sporadic and cluster outbreak area, the isolation-and-quarantine was the most cost-effective intervention. The personal protection and isolation-and-quarantine was the optimal joint strategy averting more cases than single isolation-and-quarantine. Rapid and effective isolation and quarantine could control the outbreak of COVID-19. The strategy including community containment could be more effective and cost-effective when low probability and long delay of implements of interventions or much imported cases.

## Data Availability

The data derived from previous study and they were available.

## Abbreviations

COVID-19: coronavirus disease 2019
SARS: Severe Acute Respiratory Syndrome
MERS: Middle East Respiratory Syndrome
NPIs: nonpharmaceutical interventions
ABM: agent-based model
SLIR: susceptible-latent-infectious-recovered
OR: odds ratio
RR: relative risk
CDC: Center for Disease Control and Prevention
GDP: Gross Domestic Product
PCDI: per capita disposable income
CERs: cost-effectiveness ratios
ICERs: Incremental cost-effectiveness ratios
SD: standard deviation

## Ethics approval and consent to participate

Not applicable.

## Consent for publication

Not applicable.

## Availability of data and materials

All code required to reproduce the analysis is available online.

## Authors’ Contributions

HJ and QW conceived and designed the study. HJ, QW, NYS, JXH, TTC, and LQY designed the model. QW, JA, HJ, KX, and CJB collected the parameters. QW, NYS, and JXH did the data analyses. QW, HJ, NYS, and AT contributed to the writing of the manuscript. All authors interpreted the results and approved the final version for publication.

## Acknowledgements

We would like to acknowledge editors and reviewers for their work and comments.

